# Precision shielding for COVID-19: metrics of assessment and feasibility of deployment

**DOI:** 10.1101/2020.11.01.20224147

**Authors:** John P.A. Ioannidis

## Abstract

**Background:** The ability to preferentially protect high-groups in COVID-19 is hotly debated. Here, the aim is to present simple metrics of such precision shielding of people at high-risk of death after infection by SARS-CoV-2; demonstrate how they can estimated; and examine whether precision shielding was successfully achieved in the first COVID-19 wave.

**Methods:** The shielding ratio, S, is defined as the ratio of prevalence of infection among people at a high-risk group versus among people in a low-risk group. The contrasted risk groups examined here are according to age (>=70 versus <70 years), and institutionalized (nursing home) setting. For age-related precision shielding, data were used from large seroprevalence studies with separate prevalence data for elderly versus non-elderly and with at least 1000 assessed people >=70 years old. For setting-related precision shielding, data were analyzed from 10 countries where information was available on numbers of nursing home residents, proportion of nursing home residents among COVID-19 deaths, and overall population infection fatality rate.

**Findings:** Across 17 seroprevalence studies, the shielding ratio S for elderly versus non-elderly varied between 0.4 (substantial shielding) and 1.6 (substantial inverse protection, i.e. low-risk people being protected more than high-risk people). Five studies in USA all yielded S=0.4-0.8, consistent with some shielding being achieved, while two studies in China yielded S=1.5-1.6, consistent with inverse protection. Assuming 25% infection fatality rate among nursing home residents, S values for nursing home residents ranged from 0.07 to 3.1. The best shielding was seen in South Korea (S=0.07) and modest shielding was achieved in Israel, Slovenia, Germany, and Denmark. No shielding was achieved in Hungary and Sweden. In Belgium (S=1.9), UK (S=2.2) and Spain (S=3.1), nursing home residents were far more frequently infected that the rest of the population.

**Interpretation:** The experience from the first wave of COVID-19 suggests that different locations and settings varied markedly in the extent to which they protected high-risk groups. Both effective precision shielding and detrimental inverse protection can happen in real-life circumstances. COVID-19 interventions should seek to achieve maximal precision shielding.

## RESEARCH IN CONTEXT

### Evidence before this study

There is very heated debate on whether targeted protection of high-risk groups is a feasible strategy for management of the COVID-19 pandemic. There is strong evidence for risk stratification in COVID-19 risk of death after infection, with major effects of age, institutionalized setting, and other factors. It is less known whether high-risk groups defined according to age and institutionalization criteria have been preferentially protected at all until now in the real-world experience of the first wave of COVID-19.

### Added value of this study

The shielding ratio is introduced, as the ratio of the proportion of people infected in a group at high risk of death when infected versus the group at low risk of death when infected. It is shown how the shielding ratio can be derived from seroprevalence studies; and how it can be inferred indirectly also from data on the proportion of deaths in the high-risk group. Shielding ratios are estimated for 17 large seroprevalence studies worldwide and they show wide variation (0.4-1.6), suggesting a range from substantial shielding to inverse protection (where low-risk people have been more protected than the high-risk ones). Shielding ratios are also calculated for nursing home residents in 10 countries and are found to have even greater variation (0.07-3.1). Some countries achieved precision shielding for high-risk groups according to age and institutionalization status and had minimal fatalities in the first wave. Conversely, countries with inverse protection had very high fatality loads.

### Implications of all the available evidence

Both major precision shielding and inverse protection can be seen in real-world data. Interventions to manage COVID-19 need to be examined on whether they can achieve substantial precision shielding and monitored for their impact on such shielding.

A major tension in the scientific community regarding the management of the COVID-19 pandemic is between proponents of targeted approaches, where people at high-risk are preferentially protected, and those who believe that such approaches are practically infeasible.^1-3^ The term precision shielding will be used henceforth to denote the extent to which people at higher risk of death (if infected) can be made to be less frequently infected than people for whom infection would carry a lower risk of death.

The tension between these opposing schools may be exacerbated because to-date there is mostly theoretical polarized debate without solid quantification of precision shielding. It would be useful to have standard metrics that can assess whether precision shielding is achieved. This would allow to explore the feasibility of these debated approaches in real-life data and to monitor the impact of different non-pharmaceutical interventions for COVID-19 using such metrics.

The aim of this article is to present simple metrics of precision shielding; demonstrate how they can estimated from stratified population seroprevalence data or from information on proportion of deaths occurring in high-risk groups; and examine whether precision shielding was successfully achieved in the first wave of COVID-19, or, conversely, high-risk groups were more frequently infected than low-risk groups (“inverse protection”).

## METHODS

### Conceptual background of precision shielding

Precision shielding stems from the concept of precision medicine and precision public health.^4^ The terms stratified medicine, individualized medicine, and personalized medicine are also used. The success of the concept has two prerequisites: first, the ability to identify and separate reliably individuals who have very different risks; and second, the availability of effective interventions specifically for those at high-risk. The proof that these prerequisites have been met is provided by the improved outcomes of these select, high-risk individuals who are targeted precisely.

There is very strong evidence that the risk of severe adverse outcomes and death in SARS-CoV-2 infected individuals shows extreme risk stratification according to age, and additional substantial risk stratification is possible according to gender, socioeconomic and clinical features.^5-8^ Different individuals (e.g. children versus debilitated elderly people) vary over 1000-fold in their estimated risk of death and other serious outcomes (e.g. hospitalization), if infected. Therefore, since the first prerequisite is met, the main question is whether the second prerequisite can also be met, i.e. whether interventions exist that can offer enhanced protection from SARS-CoV-2 infection targeted to those individuals who are at high-risk.

### Metrics of precision shielding

To answer this question, it is important to have some robust metrics that can assess reliably whether precision shielding is achieved or not. The most direct measure is the ratio of prevalence of ensuing infections among people at a high-risk group versus the prevalence of infections among people in a low-risk group. Let us call this ratio, S, the shielding ratio. The contrasted risk groups need to be specified: e.g. according to age (e.g. >=70 versus <70 years old), setting (e.g. institutionalized versus non-institutionalized), socioeconomic background, or multi-variable risk scores.

The potential benefit of precision shielding would be greater, when the shielding ratio is lower. A shielding ratio of S=1 means that low- and high-risk people are equally frequently infected, a shielding ratio of S=0.5 means that that high-risk people have half the risk of being infected than low-risk ones. S may also take values above 1, if somehow high-risk people get more frequently infected than low-risk people.

In this framework, the number of lives saved by precision shielding of some high-risk group is proportional to the infection fatality rate (IFR) in the high-risk group, IFR_h_, the proportion of the high-risk group in the population, f_h_, and the complement of the shielding ratio, 1-S. The number of COVID-19 deaths in the high-risk group is

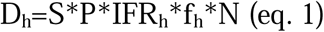

and the number of COVID-19 deaths in the low-risk group is

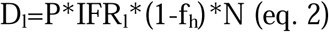

where P is the prevalence of the infection in the low-risk group and N is the total population of interest. The proportion of COVID-19 deaths occurring in the high-risk group is then given by

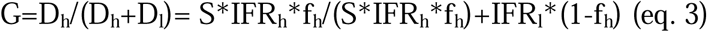

### Estimation of precision shielding from prevalence data

Seroprevalence studies that assess the frequency of infection in the general population (or in samples that try to approximate the general population) can be used to examine the extent of precision shielding achieved in different risk groups. Seroprevalence studies do have several caveats and biases that have been previously described, e.g. related to sampling and selection biases and test performance.^9,10^ When their data are used to evaluate whether precision shielding has been achieved, some additional issues should be considered. Specifically, the sampling and selection biases and the test performance of the antibody assays may be different in groups of high-versus low-risk individuals.

For example, if the high-versus low-risk groups are defined based on age, sampling bias and selection forces may be different in the two groups. E.g. individuals in poorer health (and thus at higher risk of poor outcomes if infected with SARS-CoV-2) may be less likely to be sampled. This bias may be more prominent in the elderly group in some studies or in the non-elderly group in others. Or, antibody test performance (sensitivity and specificity) may be different in the elderly versus non-elderly group, e.g. infections that do not generate detectable antibodies or where antibody titers decline faster may be more common in younger people who experience a less severe infection,^11^ or in those who are older and more debilitated to launch an immune response.

Acknowledging these caveats, one can assess the shielding ratio S for elderly (e.g. >=70 years old) versus non-elderly in large seroprevalence studies that have substantial amounts of data for both these high- and low-risk groups. Here, seroprevalence studies from 4 recent systematic reviews^10,12-14^ were screened. Studies were selected for analysis if they had at least 1000 participants >=70 years old, so that a substantial amount of data from this group would be available for a meaningful assessment against the younger age group.

Information was extracted on the crude seroprevalence in the elderly versus non-elderly group. Whenever adjusted estimates were provided (e.g. adjusting for test performance, demographics, sampling or design features, ethnicity/race, and/or other factors), the maximally adjusted estimates were also extracted for each group. Unadjusted and adjusted estimates were compared, but the latter were preferred, whenever available. The default comparison used a 70 years cut-off, but when data were not presented for this cut-off, a lower cut-off (the one most proximal to 70 years) was used. The share of the high-risk group (>=70 years) in the general population was derived from population pyramids for the respective countries/locations.

### Estimation of precision shielding from proportion of COVID-19 deaths occurring in the high-risk group

For many risk factors other than age, data on the prevalence of the infection in high- and low-risk groups may not be available. In these cases, one can estimate the shielding ratio S if the proportion of COVID-19 deaths that are contributed by the high-risk group (G), the relative share of the high-risk group in the general population (f_h_), and the IFR in the low- and high-risk groups (IFR_l_ IFR_h_) are known or can be reasonably approximated. S is then obtained by solving equation 3 for S.

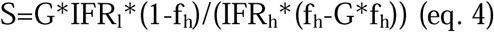

For example, extremely high risk of COVID-19 death is seen in institutionalized elderly individuals in nursing homes, where IFR_h_ is in the range of 25%,^15^ i.e. about 1000-fold higher than in non-institutionalized people. As of this writing, there are no large published seroprevalence studies to-date that have evaluated representative samples of nursing home residents at national level in different countries. However, one can estimate S by using available data from countries where there is information on the proportion of COVID-19 deaths that happened among institutionalized elderly people, the share of this group in the general population, and the IFR in the general population (and hence also in the non-institutionalized population). A range of values 15-35% for IFR_h_ may be used in sensitivity analyses with IFR=25% in the baseline scenario.

For the purposes of calculations done here, information of nursing and elderly home beds in different countries and on the proportion of COVID-19 deaths that occurred in nursing homes in the first wave is derived from the International Long Term Care Policy Network^16^ considering the last update of their review on COVID-19 mortality in nursing homes (released on October 14, 2020). Information on the overall population IFR is derived from ref. 10; data were used only from countries where IFR estimation had been informed from >1500 samples, so that there would be less uncertainty on the IFR estimate. IFR in non-institutionalized people is calculated by the overall IFR excluding the deaths in nursing homes from the numerator and the number of nursing home residents from the denominator (country population).

## RESULTS

### Precision shielding according to age

Table 1 shows the characteristics of 17 eligible large population seroprevalence studies^17-33^ where results were separately available in elderly versus younger groups. Table 2 shows the shielding ratios based on crude (unadjusted) and adjusted seroprevalence data. As shown, the shielding ratio ranged from 0.4 to 1.6.

**Table 1.** Characteristics of the eligible seroprevalence studies.

| **Location (ref)** | **Type of population** | **Date** | **Sample tested, n** | **Age cutoff,\* y** |
| --- | --- | --- | --- | --- |
| EUROPE |  |  |  |  |
| Denmark (17) | Blood donors | 1-26 June | 2311 | 70 |
| Hungary (18) | General | 1-16 May | 10472 | 65 |
| Iceland (19) | General | April-June | 30576 | 70 |
| Spain (20) | General | 14 April – 1 May | 61075 | 65 |
| UK (21) | General | 20 June-13 July | 99908 | 65 |
| UK (22) | Biobank | 27 May-14 August | 18734 | 70 |
| AMERICAS |  |  |  |  |
| Brazil (23) | General | 4-7 June | 31165 | 70 |
| Canada (24) | Blood donors | May | 37737 | 60 |
| Dominican Republic(25) | General, hotspot areas | April-June | 12897 | 60 |
| USA (26) | Hemodialysis | July | 28503 | 65 |
| USA (27) | Life insurance applicants | 12 May – 25 June | 50025 | 61 |
| USA (28) | Blood donors | June-July | 189656 | 65 |
| USA-New York (29) | General, convenience | 19-28 April | 15101 | 55 |
| USA-Brooklyn (30) | General, convenience | Early May | 11092 | 70 |
| ASIA |  |  |  |  |
| China -Wuchang (31) | General | Mid-May | 61437 | 70 |
| China (32) | Diverse | January-April | 17368 | 65 |
| India (33) | General | 11 May- 4 June | 28000 | 60 |

\*for the comparison of seroprevalence in elderly versus younger participants, 70 years was the default cut-off chosen, but as shown in this column, a lower cut-off was chosen when seroprevalence data according to the 70 year cut-off were not available

**Table 2.** Estimates of the shielding ratio for elderly people versus younger people in different locations for the first wave of COVID-19

| Location (ref) | Prevalence (%) |  | S (95% CI) | Adjusted prevalence (%) |  | S (95% CI) | f<sub>h</sub> (%) |
| --- | --- | --- | --- | --- | --- | --- | --- |
|  | Elderly | Younger |  | Elderly | Younger |  |  |
| EUROPE |  |  |  |  |  |  |  |
| Denmark (17) | 1.8 | 3.0 | 0.6 (0.4-1.1) | 1.4 | 2.5 | 0.6 (0.3-1.1) | 14 |
| Hungary (18) | 0.8 | 0.6 | 1.3 (0.7-2.1) | ND | ND | ND | 12 |
| Iceland (19) | 0.5 | 1.0 | 0.5 (0.4-0.6) | ND | ND | ND | 10 |
| Spain (20) | ND | ND | ND | 6.0 | 4.7 | 1.3 (1.2-1.4) | 15 |
| UK (21) | 2.7 | 5.6 | 0.5 (0.4-0.6) | 3.3 | 6.7 | 0.5 (0.4-0.6) | 14 |
| UK (22) | 6.1 | 8.8 | 0.7 (0.6-0.8) | ND | ND | ND | 14 |
| AMERICAS |  |  |  |  |  |  |  |
| Brazil (23) | 2.4 | 2.8 | 0.9 (0.7-1.1) | 2.4 | 2.8 | 0.8 (0.6-1.2) | 6 |
| Canada (24) | 0.7 | 0.7 | 1.0 (0.7-1.3) | ND | ND | ND | 12 |
| Dominican Republic (25) | 6.0 | 5.3 | 1.1 (*) | ND | ND | ND | 5 |
| USA (26) | 7.6 | 8.4 | 0.9 (0.8-1.0) | 8.1 | 9.7 | 0.8 (0.8-0.9) | 11 |
| USA (27) | 2.0 | 3.1 | 0.6 (0.5-0.8) | ND | ND | ND | 11 |
| USA (28) | 0.8 | 1.8 | 0.4 (0.4-0.5) | ND | ND | ND | 11 |
| USA-New York (29) | 10.9 | 13.5 | 0.9 (0.8-1.0) | 12.1 | 15.4 | 0.8 (0.7-0.9) | 10 |
| USA-Brooklyn (30) | ~40 | ~50 | 0.8 (*) | ND | ND | ND | 9 |
| ASIA |  |  |  |  |  |  |  |
| China - Wuchang (31) | 3.5 | 2.2 | 1.6 (1.4-1.8) | ND | ND | ND | 7 |
| China (32f) | 2.0 | 1.3 | 1.5 (1.2-1.9) | ND | ND | ND | 7 |
| India (33) | 0.6 | 0.6 | 1.0 (0.7-1.5) | ND | ND | ND | 4 |

\* information available does not allow reliable calculation of 95% confidence intervals
The exact number of participants $\geq 70$ years tested is not provided for the studies in Iceland, Canada and Dominican Republic, but based on the number of participants in the highest provided age stratum and the age-structure of the population in these countries, it is likely that those $\geq 70$ would exceed the minimum required sample of $n=1000$ . For Iceland, the data include the cases detected with positive PRC test (the majority) plus those estimated to be infected based on antibody testing.
ND, no data; S, shielding ratio; CI, confidence interval; f<sub>h</sub>, population $\geq 70$ years old in the location.

The elderly were infected substantially more frequently than the younger populations in Spain and China and possibly in Hungary, although in Hungary the 95% confidence interval (CI) could not exclude values less than 1. Two studies in China yielded very similar estimates of S (1.5-1.6). The Dominican Republic, India, and Canada had S values very close to 1.0 (no shielding) and 95% CIs exclude major shielding. Modest shielding was suggested in Brazil (estimated S=0.8), but the 95% CI could not exclude values above 1. In the USA, five studies all show S value estimates below 1, with three of them yielding modest estimates of S=0.8 and the other two yielding S=0.6 and S=0.4, but with largely overlapping CIs. The two lowest estimates come from studies on life insurance applicants and blood donors, where selection biases that depend on age cannot be excluded. E.g. stronger healthy volunteer bias may exist in the elderly life insurance applicants and blood donors. The best S values (S=0.5-0.7) suggesting relatively successful age-related shielding were seen in Denmark, Iceland and in 2 studies in the UK, but the 95% CI had large uncertainty in Denmark.

Of note, most of the seroprevalence estimates in Table 1 were from crude, unadjusted analyses (positive samples per total tested samples). Adjustments were used only in the Denmark, Spain, UK, Brazil, USA-hemodialysis, USA-New York studies, and adjusted point estimates tended to be similar to the unadjusted ones. The proportion of people >=70 years old in the general population was 9-15% in high-income countries, and 4-7% in other countries.

### Precision shielding according to institutionalized setting

Table 3 runs calculations for precision shielding of nursing home residents in 10 countries during the first wave of COVID-19. They include 8 European countries, Israel and South Korea.

**Table 3.** Estimating the shielding ratio for institutionalized nursing home residents versus the rest of the population in 10 countries during the first wave of COVID-19

| Country | Nursing home residents per 100,000 population | Proportion COVID-19 deaths in nursing homes, G (%) | Overall population IFR (%) | IFR in non-institutionalized, IFR<sub>1</sub> (%) | Shielding ratio, S |
| --- | --- | --- | --- | --- | --- |
| Belgium | 1080 | 61 | 0.87 | 0.34 | 1.9 (1.4-3.2) |
| Denmark | 690 | 35 | 0.27 | 0.18 | 0.5 (0.4-0.9) |
| Germany | 980 | 39 | 0.23 | 0.14 | 0.4 (0.3-0.6) |
| Hungary | 570 | 23 | 0.54 | 0.42 | 0.9 (0.6-1.4) |
| Slovenia | 1100 | 81 | 0.11 | 0.02 | 0.3 (0.2-0.5) |
| Spain | 690 | 63 | 0.85 | 0.31 | 3.1 (2.2-5.1) |
| Sweden | 810 | 46 | 0.57 | 0.31 | 1.3 (0.9-2.1) |
| UK (England) | 760 | 45 | 0.93 | 0.51 | 2.2 (1.6-3.7) |
| South Korea | 420 | 8 | 0.09 | 0.08 | 0.07 (0.05-0.11) |
| Israel | 520 | 39 | 0.10 | 0.06 | 0.3 (0.2-0.5) |

IFR, infection fatality rate
S, shielding ratio. S is calculated from equation 4 assuming IFR in nursing home residents of 25% in the baseline scenario and 15% or 35% in the scenarios of sensitivity analyses that define the presented bounds.

As shown, Belgium, UK, and Spain have had very unfavorable S values (S=1.9, 2.2. and 3.1 in the baseline scenario, potentially even higher in sensitivity analyses). This means that people who were institutionalized in nursing homes were approximately twice as likely to be infected than the non-institutionalized population in Belgium and the UK, and more than 3 times more likely to be infected than the non-institutionalized population in Spain.

Sweden and Hungary seemed to have been unable to protect their nursing homes more than the general population and sensitivity analysis for Sweden suggests that it is possible that nursing home residents were even infected more than twice as frequently than the rest of the population.Some substantial shielding was seen in Denmark, Slovenia, Germany, and Israel (S=0.3-0.5 in the baseline scenario). Extremely effective shielding was achieved in South Korea, where one can estimate S=0.07. Only 8% of COVID-19 deaths in this country occurred in nursing home residents.

## DISCUSSION

### Shielding ratio and overall load of fatalities

The shielding ratio can be used as a metric to assess whether protection of high-risk populations is being achieved in a given country or jurisdiction. As shown, data from the first wave of COVID-19 suggest that the shielding ratio can take very different values, ranging from extremely effective protection of vulnerable high-risk populations to major inverse protection, where high-risk populations have been protected far less successfully than low-risk populations.

Fatality rates tend to be relatively low in countries where the elderly (and even more so the institutionalized elderly) have been effectively protected. It is possible that one can achieve better values of shielding (lower S) in nursing homes than in non-institutionalized elderly who are unavoidably more freely mobile in the community. Countries that have avoided massive infections in nursing homes, have had much lower fatality burden from COVID-19 in the first wave. It is estimated^16^ that in the first wave only 0.01% of South Korean nursing home residents died with COVID-19, as opposed to 3.3% in Sweden and more than 5% in Belgium, England, and Spain. While there may be differences on how deaths are attributed to COVID-19 among nursing home residents, these are unlikely to explain away such major differences across countries. Besides nursing homes, some differential protection can be achieved even for the non-institutionalized elderly and this may result in substantially lower fatalities overall. Thus, Iceland and Denmark did have 20% and 35% of the COVID-19 occur in nursing homes, but they seem to have protected effectively their elderly; therefore, they have had low fatalities in the first wave.

The worst fatality rates have been seen in locations with high proportions of elderly and/or institutionalized people and where there was strong inverse protection. For example, Castiglione d’Adda,^34^ a small town in Lombardy had 47 COVID-19 deaths in a population of 4550 people. Seroprevalence data^34^ showed IgG positivity in 51/155 people >=60 years old versus 64/290 in younger people, which translate into S=1.5 for age-related shielding and the town also had nursing homes affected. Another seroprevalence study in Northern Italy locations found that seroprevalence was 4.5 times larger in nursing home residents compared to non-institutionalized people.^35^ While these data may not necessarily be representative of Italy as a whole, they are congruent with the very high fatalities in particular areas of Lombardy.^36^

Some countries may have had mixed patterns, e.g. protecting somehow their elderly, but not specifically their institutionalized elderly, as in the case of the UK and probably also the USA where 44% of COVID-19 deaths occurred in the 0.59% of the population that resides in nursing homes.^16^ This pattern can still translate to heavy cumulative death toll. Institutionalized elderly are at much higher risk of death than other elderly people, and they can contribute a lion’s share to the overall death count.

While only age and nursing home residence were explored here, other risk factors may also be assessed in a similar fashion in terms of the extent of precision shielding. For example, socioeconomic factors are known to be strong determinants of the infection rate.^37^ Minorities and disadvantaged populations are more likely to be infected and it is possible that may also have more adverse outcomes due to poorer health status.

### Impact of horizontal and targeted measures on precision shielding metrics

Different measures against the spread of COVID-19 need to be assessed in terms of their effect on precision shielding. One might argue that horizontal measures to mitigate COVID-19 for everyone without making discriminations according to risk would have S=1, as infection rates would be decreased equally in all groups. In many/most circumstances, this may not hold true. Most measures may eventually leave some population subgroups more exposed than others. The groups that still remain unavoidably highly exposed may occasionally be among those that have lower risk (e.g. young, healthy military personnel in congested areas like barracks or military vessels). However, in most situations horizontal measures may unintentionally leave high-risk groups more exposed than low-risk groups.

For example, horizontal lockdown measures typically protect young, healthy professionals who can work from home, but leave far more exposed the essential workers and those who are disadvantaged, e.g. the homeless. These poorly sheltered populations often have a higher burden of background comorbidities and more limited medical care – and are thus at higher risk of death, if infected by SARS-CoV-2. Similarly, horizontal lockdowns may leave nursing home populations less protected than non-institutionalized populations, unless additional targeted measures are taken focused on nursing homes specifically. Nursing home residents have very limited mobility and often live together in closed, congested spaces – as opposed to young, healthy individuals who shelter in place alone or in smaller numbers with their families. Thus, massive infections are easier to occur in nursing homes. The situation may become even worse, if nursing home personnel also has a high S value, since personnel will then infect the residents. This was apparently the case with Stockholm during the first wave, where seroprevalence among nursing home personnel was 23% in the first 20 days of April,^38^ three times higher than general population of Stockholm at the same time. Nursing home personnel in Stockholm was highly mobile and exposed across different nursing homes.

Lockdown measures also force young low-risk individuals to spend more time indoors and this may increase the exposure of any high-risk family members who have to live in the same house.

### Implications for future interventions and metrics of their success

The most-widely used metric for the success of interventions against COVID-19 to-date has been the number of infections. This metric alone is problematic, because the vast majority of infections remain unrecorded and the documented infections depend on how many tests are done. A more informative metric of success is the ability of different interventions to generate a most favorable shielding ratio for the most high-risk subgroups of the population. These subgroups may account for the vast majority of the potential deaths and, if properly protected, many deaths can be avoided.

Estimates for the shielding ratio and its evolution as the epidemic wave progresses and as different interventions are employed can be obtained from prevalence studies using antibodies, or other tests (e.g. antigen testing).^39,40^ Alternatively, they can be obtained from assessments of the profile of fatalities, provided that the relative representation of the high-risk groups of interest in the general population is known, and that some fair estimate of the IFR in low-and high-risk groups exists from previous studies. S can be used as an outcome in COVID-19 interventional studies.^41^

## Limitations

Estimates of precision shielding may have some considerable error margin and biases, inherent in the parameters that go in the calculation of S. Thus, one has to interpret the ensuing S values with caution and allow for substantial uncertainty. These values should be able to convey whether some substantial shielding is achieved, or whether gross inverse protection is making things worse. Small differences in S estimates should not be over-interpreted. Validation with multiple, preferably large and unbiased, studies on the same population may help get a better sense of the accuracy and heterogeneity of such estimates.

Most data analyzed here came from high income countries. Limited data from two other countries (India and Dominican Republic) showed estimates of S very close to 1, suggesting no achieved shielding. It is possible that shielding is more difficult to achieve in some resource-poor settings and in congested, highly-mixing populations where most people cannot shelter effectively and have limited private space, living in multigenerational families. Similarly, even within the same country, or sub-location, there may be sub-populations that can achieve much better precision shielding than others, due to socioeconomic circumstances and other factors. Job circumstances may be particularly important and further studies should evaluate precision shielding in different high-risk jobs. For example, some data suggest that age-specific precision shielding can be successfully achieved for health care, first response, and public safety personnel: S was 0.4 for such personnel based on a 65 years cut-off in Detroit.^42^

### Precision health and COVID-19

Precision approaches had received enthusiastic support before the COVID-19 era as a way to transform medicine and health at large. Hopes (and hype)^43,44^ were fueled in particular by perceived improvements in predictive ability, especially with the advent of -omics. However, the discriminating ability of most -omics discoveries had been modest: e.g. single gene variants often differentiated risk by less than 1.1-fold and even complex molecular signatures and multi-gene models often differentiated risk by less than 3-fold between high- and low-risk individuals. In this regard, COVID-19 offers a situation where risk discrimination is far better than most previous efforts at materializing precision medicine. If the risk stratification offered by COVID-19 does not suffice for precision purposes, then it is unlikely that the concept of precision medicine can find fruitful applications with major impact across medicine (perhaps with the exception of some rare conditions). At a minimum, it is worth trying to make precision approaches work for COVID-19. Even modest shielding ratios may translate into hundreds of thousands or even millions of lives saved during the multi-year course of the pandemic.^45^

## Data Availability

All the relevant data are in the paper

## Conflicts of interest

I have no conflicts of interest. I have received no funding for my COVID-19 work. I have not signed the Great Barrington Declaration or the John Snow Memorandum or any other petitions that resolve around the discussed issues. METRICS has been funded by grants from the Laura and John Arnold Foundation.

## Funding

none

## Notes

### Competing Interest Statement

The authors have declared no competing interest.

### Funding Statement

No funding was received for this work.

### Author Declarations

No new data were collected; modeling of existing data

## References

1. Smith GD. Shielding from covid-19 should be stratified by risk. BMJ 2020;369:m2063

2. Great Barrington Declaration, in: https://gbdeclaration.org/, xlast accessed November 1, 2020.

3. John Snow Memorandum, in: https://www.johnsnowmemo.com/, xlast accessed November 1, 2020.

4. Khoury MJ, Iademarco MF, Riley WT. Precision public health for the era of precision medicine. Am J Prev Med. 2016;50(3):398–401.

5. Spiegelhalter D. Use of “normal” risk to improve understanding of dangers of covid-19. BMJ 2020;370:m3259.

6. Ioannidis JPA, Axfors C, Contopoulos-Ioannidis DG. Population-level COVID-19 mortality risk for non-elderly individuals overall and for non-elderly individuals without underlying diseases in pandemic epicenters. Environ Res. 2020 Sep;188:109890.

7. Williamson EJ, Walker AJ, Bhaskaran K, Bacon S, Bates C, Morton CE, Curtis HJ, Mehrkar A, Evans D, Inglesby P, Cockburn J, McDonald HI, MacKenna B, Tomlinson L, Douglas IJ, Rentsch CT, Mathur R, Wong AYS, Grieve R, Harrison D, Forbes H, Schultze A, Croker R, Parry J, Hester F, Harper S, Perera R, Evans SJW, Smeeth L, Goldacre B. Factors associated with COVID-19-related death using OpenSAFELY. Nature. 2020;584(7821):430–436.

8. Grant A. Apparent reductions in COVID-19 Case Fatality Rates reflect changes in average age of those testing positive. medRxiv 2020; https://doi.org/10.1101/2020.09.18.20197160

9. Ioannidis JPA. Infection fatality rate of COVID-19 inferred from seroprevalence data. Bulletin WHO 2020; https://www.who.int/bulletin/online_first/BLT.20.265892.pdf.

10. Burgess S, Ponsford MJ, Gill D. Are we underestimating seroprevalence of SARS-CoV-2? BMJ. 2020;370:m3364.

11. Seow J, Graham C, Merrick B, Acors S, Steel KJA, Hemmings O, et al. Longitudinal evaluation and decline of antibody responses in SARS-CoV-2 infection [preprint]. Cold Spring Harbor: medRxiv; 2020. doi: https://doi.org/10.1101/2020.07.09.20148429 doi: http://dx.doi.org/10.1101/2020.07.09.20148429

12. Franceschi VB, Santos AS, Glaeser AB, et al. Population-based prevalence surveys during the COVID-19 pandemic: a systematic review. medRxiv 2020; doi: https://doi.org/10.1101/2020.10.20.20216259

13. Chen X, Chen Z, Azman AS, et al. Serological evidence of human infection 1 with SARS-CoV-2: a systematic review and meta-analysis. medRxiv 2020; doi: https://doi.org/10.1101/2020.09.11.20192773

14. Arora RK, Joseph A, Van Wyk J, Rocco S, Atmaja A, May E, Yan T, Bobrovitz N, Chevrier J, Cheng MP, Williamson T, Buckeridge DL. SeroTracker: a global SARS-CoV-2 seroprevalence dashboard. Lancet Infect Dis. 2020;S1473-3099(20)30631-9.

15. Arons MM, Hatfield KM, Reddy SC, et al. Presymptomatic SARS-CoV-2 infections and transmission in a skilled nursing facility. N Engl J Med. 2020;382(22):2081–2090.

16. International Long Term Care Policy Network. Mortality associated with COVID-19 in care homes: international evidence. In: https://ltccovid.org/2020/04/12/mortality-associated-with-covid-19-outbreaks-in-care-homes-early-international-evidence/#:~:text=Based%20on%20the%20data%20gathered,(based%20on%2021%20countries, last accessed November 1, 2020.

17. Pederson OB, Nissen J, Dinh KM, et al. SARS-CoV-2 infection fatality rate among elderly retired Danish blood donors - A cross-sectional study. Clin Infect Dis 2020 Oct 26;ciaa1627. doi: 10.1093/cid/ciaa1627.

18. Merkely B, Szabó AJ, Kosztin A, Berényi E, Sebestyén A, Lengyel C, et al.; HUNgarian COronaVirus-19 Epidemiological Research (H-UNCOVER) investigators. Novel coronavirus epidemic in the Hungarian population, a cross-sectional nationwide survey to support the exit policy in Hungary. Geroscience. 2020;42(4):1063–74.

19. Gudbjartsson DF, Norddahl GL, Melsted P, Gunnarsdottir K, Holm H, Eythorsson E, et al. Humoral immune response to SARS-CoV-2 in Iceland. N Engl J Med. 2020 Sep 1.NEJMoa2026116.

20. Pollán M, Pérez-Gómez B, Pastor-Barriuso R, Oteo J, Hernán MA, Pérez-Olmeda M, et al.; ENE-COVID Study Group. Prevalence of SARS-CoV-2 in Spain (ENE-COVID): a nationwide, population-based seroepidemiological study. Lancet. 2020;396(10250):535–44.

21. Ward H, Atchinson C, Whitaker M, Ainslie KED, Elliott J, Okell L, et al. Antibody prevalence for SARS-CoV-2 following the peak of the pandemic in England: REACT2 study in 100,000 adults [preprint]. Cold Spring Harbor: medRxiv; 2020. doi: https://doi.org/10.1101/2020.08.12.20173690 doi: http://dx.doi.org/10.1101/2020.08.12.20173690

22. UK Biobank. UK Biobank SARS-CoV-2 serology study, September 16, 2020.

23. Hallal PC, Hartwig FP, Horta BL, et al. SARS-CoV-2 antibody prevalence in Brazil: results from two successive nationwide serological household surveys. Lancet Glob Health 2020;8:e1390–98.

24. Canadian Blood Services. COVID-19 Seroprevalence Report – August 19, 2020.

25. Paulino-Ramirez R, Ba’ez AA, Degaudenzi AV, Tapia L. Seroprevalence of specific antibodies against SARS-CoV-2 from hotspot communities in the Dominican Republic. Am J Trop Med Hyg. 2020, doi:10.4269/ajtmh.20-0907.

26. Anand S, Montez-Rath M, Han J, Bozeman J, Kerschmann R, Beyer P, Parsonnet J, Chertow GM. Prevalence of SARS-CoV-2 antibodies in a large nationwide sample of patients on dialysis in the USA: a cross-sectional study. Lancet 2020;396:1335–44.

27. Rigatti SJ, Stout R. SARS-CoV-2 antibody prevalence and association with routine laboratory values in a life insurance applicant population. medRxiv 2020; https://doi.org/10.1101/2020.09.09.20191296.

28. Vassallo RR, Bravo MD, Dumont LJ, Hazegh K, Kamel H. Seroprevalence of antibodies to SARS-CoV-2 in US blood donors. medRxiv 2020; https://doi.org/10.1101/2020.09.17.20195131.

29. Rosenberg ES, Tesoriero JM, Rosenthal EM, Chung R, Barranco MA, Styer LM, et al. Cumulative incidence and diagnosis of SARS-CoV-2 infection in New York. Ann Epidemiol. 2020 Aug;48:23–29.e4.

30. Reifer J, Hayum N, Heszkel B, Klagsbald I, Streva VA. SARS-CoV-2 IgG antibody responses in New York City [preprint]. Cold Spring Harbor: medRxiv; 2020. doi: https://doi.org/10.1101/2020.05.23.20111427 doi: http://dx.doi.org/10.1101/2020.05.23.20111427

31. Pan Y, Li X, Yang G, et al. Seroprevalence of SARS-CoV-2 immunoglobulin antibodies in Wuhan, China: part of the city-wide massive testing campaign. Clin Microbiol Infect 2020;

32. Xu X, Sun J, Nie S, Li H, Kong Y, Liang M, et al. Seroprevalence of immunoglobulin M and G antibodies against SARS-CoV-2 in China. Nat Med. 2020 08;26(8):1193–5.

33. Murhekar M, Bhatnagar T, Selvaraju S, Rade K. Prevalence of SARS-CoV-2 infection in India: Findings from the national serosurvey, May-June 2020. Indian J Med Res 2020;.

34. Pagani G, Conti F, Giacomelli A, et al. Seroprevalence of SARS-CoV-2 significantly varies with age: Preliminary results from a mass population screening. J Infect 2020; https://doi.org/10.1016/j.jinf.2020.09.021.

35. Vena A, Berruti M, Adessi A, et al. Prevalence of antibodies to SARS-CoV-2 in Italian adults and associated risk factors. J Clin Med 2020;9:2780.

36. Boccia S, Ricciardi W, Ioannidis JPA. What other countries can learn from Italy during the COVID-19 pandemic. JAMA Intern Med. 2020 Jul 1;180(7):927–928.

37. Iacobucci G. Covid-19: Increased risk among ethnic minorities is largely due to poverty and social disparities, review finds. BMJ 2020; DOI: 10.1136/bmj.m4099.

38. Lindahl JF, Hoffman T, Esmaeilzadeh M, et al. High seroprevalence of SARS-CoV-2 in elderly care employees in Sweden. Infect Ecology Epidemiol 2020;10:1789036, DOI: 10.1080/20008686.2020.1789036.

39. Mak GC, Cheng PK, Lau SS, Wong KK, Lau CS, Lam ET, Chan RC, Tsang DN. Evaluation of rapid antigen test for detection of SARS-CoV-2 virus. J Clin Virol. 2020 Aug;129:104500.

40. Nagura-Ikeda M, Imai K, Tabata S, Miyoshi K, Murahara N, Mizuno T, Horiuchi M, Kato K, Imoto Y, Iwata M, Mimura S, Ito T, Tamura K, Kato Y. Clinical Evaluation of Self-Collected Saliva by Quantitative Reverse Transcription-PCR (RT-qPCR), Direct RT-qPCR, Reverse Transcription-Loop-Mediated Isothermal Amplification, and a Rapid Antigen Test To Diagnose COVID-19. J Clin Microbiol. 2020 Aug 24;58(9):e01438–20.

41. Cristea IA, Naudet F, Ioannidis JPA. Preserving equipoise and performing randomized trials for COVID-19 social distancing interventions. Epidemiol Psychiatr Sci. 2020 Oct 28:1–27. doi: 10.1017/S2045796020000992.

42. Akinbami LJ, Vuong N, Petersen LR, Sami S, Patel A, Lukacs SL, Mackey L, Grohskopf LA, Shehu A, Atas J. SARS-CoV-2 Seroprevalence among Healthcare, First Response, and Public Safety Personnel, Detroit Metropolitan Area, Michigan, USA, May–June 2020. Emerg Infect Dis 2020; 2020 Sep 21;26(12). doi: 10.3201/eid2612.203764; https://doi.org/10.3201/eid2612.203764.

43. Collins FS, Varmus H. A new initiative on precision medicine. N Engl J Med 2015;372:793–795.

44. Bossuyt PMM. The thin line between hope and hype in biomarker research. JAMA. 2011;305(21):2229–2230. doi:10.1001/jama.2011.729

45. Ioannidis JPA. Global perspective of COVID-19 epidemiology for a full-cycle pandemic. Eur J Clin Invest. 2020 Oct 7;e13421.

